# Post-pandemic Spread of Influenza and RSV in a Child Care Facility in Ontario in the Presence of Vaccination

**DOI:** 10.1101/2025.11.24.25340890

**Authors:** D. Flynn-Primrose, B. Amoako, L. Humphrey, Z. Mohammadi, E. W. Thommes, J. Lee, D. Neame, M. G. Cojocaru

## Abstract

We present a mathematical model of a daycare center in Ontario to investigate concurrent pathogen transmissions among students (1.5 to 4 years) and teachers in a simulated child care center. The pathogens included are seasonal influenza and RSV. The model simulates detailed movements and interactions within a structured childcare environment, enabling analysis of pathogen exposure, infection rates, and transmission dynamics. Simulations incorporate empirical contact data collected from an Ontario daycare facility, existing values of epidemiological parameters, and publicly available health statistics. Pathogens are assumed to be introduced from outside of the facility, and the model provides estimates of infection rates within the childcare facility, as well as likelihood of re-introduction from offsite. Results derived from the model further explore the impact of existing and potential vaccination strategies on diseases transmission. We explore scenarios reflecting current vaccination uptake for influenza, as well as potential uptake and efficacy of a novel RSV vaccine.

## 1. Introduction

### 1.1. Literature Review

Agent-based modeling has emerged as a valuable tool for understanding the dynamics of infectious disease spread. These models simulate the behavior of individuals (agents) within a population, allowing researchers to capture characteristics and behaviors that impact the complexities of disease transmission and intervention strategies. Models often incorporate real-world data, such as age groups, social networks, mobility patterns, and environmental factors, to create realistic scenarios. In [1], Adalja et al investigated the effects of social contact networks on pathogen dissemination in a setting analogous to a daycare center or grade school. They showed that interventions that decrease mixing within child care facilities could diminish pathogen dissemination and possibly amplify the effectiveness of vaccination. Chumachenko et al [2] demonstrate how to embed analytic SEIR equations within an agent-based framework, validate the approach qualitatively against known epidemic patterns of influenza and other acute respiratory viral infections (ARVI), and argue that such hybrid models can offer public-health planners more flexibility than purely differential-equation or statistical tools.

Lang’s 2022 systematic review identified barely a dozen individual-based RSV models worldwide, most focused on households rather than institutional venues [3]. One of these papers [4] used Kenyan cohort data to show that symptomatic individuals are up to seven times more infectious than asymptomatic individuals, especially those with a high viral load, and half of RSV spread occurs within households. An individual-based simulation model of RSV transmission dynamics with vaccination in 13 high-income countries projects that vaccinating older adults (above 60 years) with the prefusion F (RSVpreF) vaccine which is available for older adults could avert 35–64% of hospitalizations within the age group, while maternal vaccination could prevent up to 50% of infant admissions [5].

### 1.2. Data for Model Parametrization

In this paper, we analyze the concurrent spread of two pathogens in a child care center, although the model allows for any other number of concurrent pathogens the modeler wishes to test. Influenza and respiratory syncytial virus (RSV) have, post-pandemic, proven to be extremely active [6], [7]. In order to analyze the spread of these pathogens, beyond monitoring agents’ behavior, it was essential to understand the key clinical features of infections from current literature analysis [8], [9]. In particular, information on the onset and progression of symptoms was essential for incorporating realistic illness-related absences into the simulated learning center. Data from the Canadian Labour Force Survey [10] were used to predict absenteeism in the simulated center in both students and teachers [11].

Results from the TransFLU influenza transmission study from the National Library of Medicine allowed for secondary attack rates from asymptomatic and symptomatic influenza virus shedders to be understood, allowing for better estimation of transmission rates [12]. Further information from the National Library of Medicine allowed simulations to be adjusted to match the burden of disease of influenza in Canada [13]. Lastly, it was essential to understand viral shedding and transmission in both symptomatic and asymptomatic agents [14].

It was equally important to understand the properties of RSV transmission in a population where age, the severity of the infection, and co-infection have an impact on the duration of RSV shedding [15]. The Centers for Disease Control and Prevention provided information regarding the transmission of RSV, such as the transmission ratios upon contact between infected and uninfected agents and the amount of time between exposure periods and the beginning of the infected period [9]. Given existing RSV vaccines available to children up to the age of 24 months [16], it was necessary to know the results of tests regarding RSV infection in children in that age group [17].

Analysis of the 2023-24 seasonal influenza vaccination data from the Government of Canada [18] allowed simulations to recreate vaccine uptake levels in all agents between October and November to monitor the transmission of pathogens between agents with varying transmission values between vaccinated and unvaccinated agents. To accurately estimate the vaccine uptake levels, it was essential to have information on childcare workers in Canada, such as age and gender ratios between workers [19].

### 1.3. Small-Scale-High-Resolution ABM Model of a Child Care Center

In a previous paper [20], the authors describe a small-scale-high-resolution agent-based model of a single classroom in a child care facility. The classroom is represented in the simulation as a rectangular graph of fixed dimension containing a lattice of nodes calibrated by children step-size (gait) using blueprints of the University of Guelph Child Care Center. Every node in the graph is connected to its eight immediate neighbours (excluding nodes on the edge of the graph which have fewer neighbours). Every agent in the simulation is represented as occupying a single node in the graph at any given time, with agents being considered in contact with each other if they occupied neighbouring nodes for a sufficiently long period of time. This model explicitly simulated the movements of children and teachers over 15-minute periods between 7am to 5pm daily, for a total period of 1 week, in a simulated classroom by assigning teachers and students different movement rules summarized in Table 1. The simulation included several parameters governing which movement rules each agent followed and how energetically they followed said rule.

**Table 1.** Movement patterns for different agent type over time.

|  | Random | Following | Gathering |
| --- | --- | --- | --- |
| Children | Agent has an equal chance of staying in place or moving to an adjacent node. | Agent chooses another agent at random and minimizes their distance to that agent. | When instructed, agent will minimize distance to the relevant teacher. They continue in this pattern until dismissed. |
| Teachers | Agent may move up to two nodes in any direction. This only occurs if the agent is in a room with no students. | Agent chooses a child at random and minimizes their distance to that child. | Agent selects three children at random and instructs them to gather. |

The research team had access to empirically observed data regarding teacher and student movements collected from the Child Care Learning Center (CCLC) at the University of Guelph. The artificial contact matrices generated by the simulation were compared against the genuine contact matrices collected by observation using the Mann-Whitney-Wilcoxon test. This allowed us to calibrate the model parameters so as to synthetically generate realistic contact matrices for a variety of scenarios. These contact matrices were saved in a large library for a variety of classroom profiles, as a classroom could contain toddler or preschool children, each respectively with a maximum capacity of 10 students (toddler) to 16-24 students (preschool), and teacher-to-child ratios, respectively of 1:5 and 1:8 (see [20]). We will show in this paper that, by simulating several such classrooms simultaneously, as well as washrooms and a teachers lounge, we are able to simulate a complete child care facility. In particular we use this expanded model to investigate the spread of Influenza and RSV throughout the facility.

## 2. Concurrent Pathogens Transmission with SEIRV Model

The simulation described in section 1 is relatively limited both because it is confined to a single room and because it only models contacts between agents and does not include any pathogens to be transmitted. Our work here builds on that result to create an agent-based model of an entire childcare facility, capable of tracking the spread of multiple pathogens simultaneously. This is accomplished by treating the facility as a collection of rooms with agents moving between different rooms in the facility, according to a fixed schedule based on that used by the Child Care and Learning Centre where the empirical contact data was originally collected. Each simulated day is divided into 40 15-min periods with the state of the model at any given time being represented by a single data frame: each row in the data frame represents an agent and each column represents a piece of information about that agent. We assume that every agent remains in a single classroom throughout each 15 minute period, with the arrival and departure of agents from the facility, as well as the movement of agents between rooms, occurring in between these periods according to a fixed schedule. This allows us to use our previous results from [20] to generate separate contact matrices for each room in the facility which lets us simulate the transmission of pathogens.

There are a minimum of ten columns in the state data frame as well as six additional columns for every pathogen being simulated. The default columns are described in Table 2.

**Table 2.**
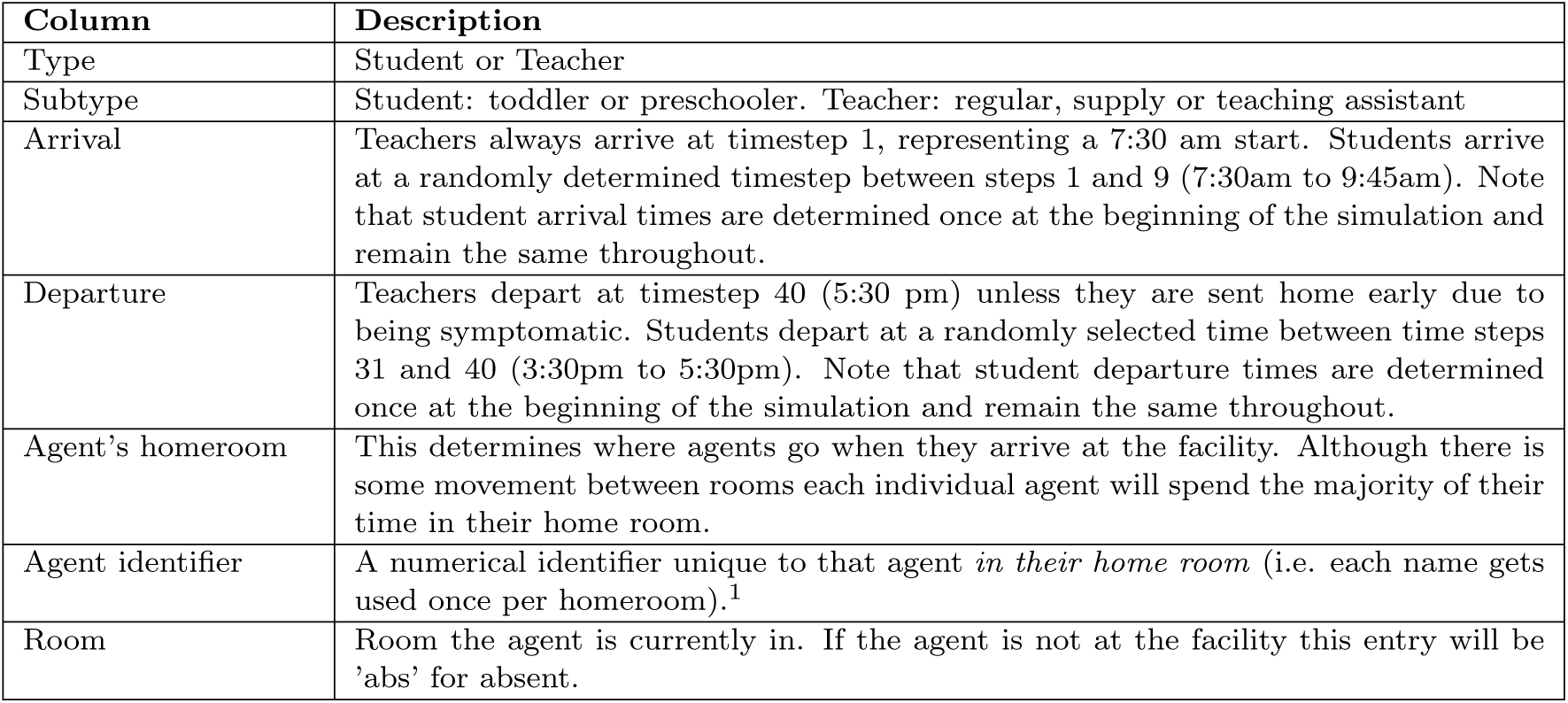
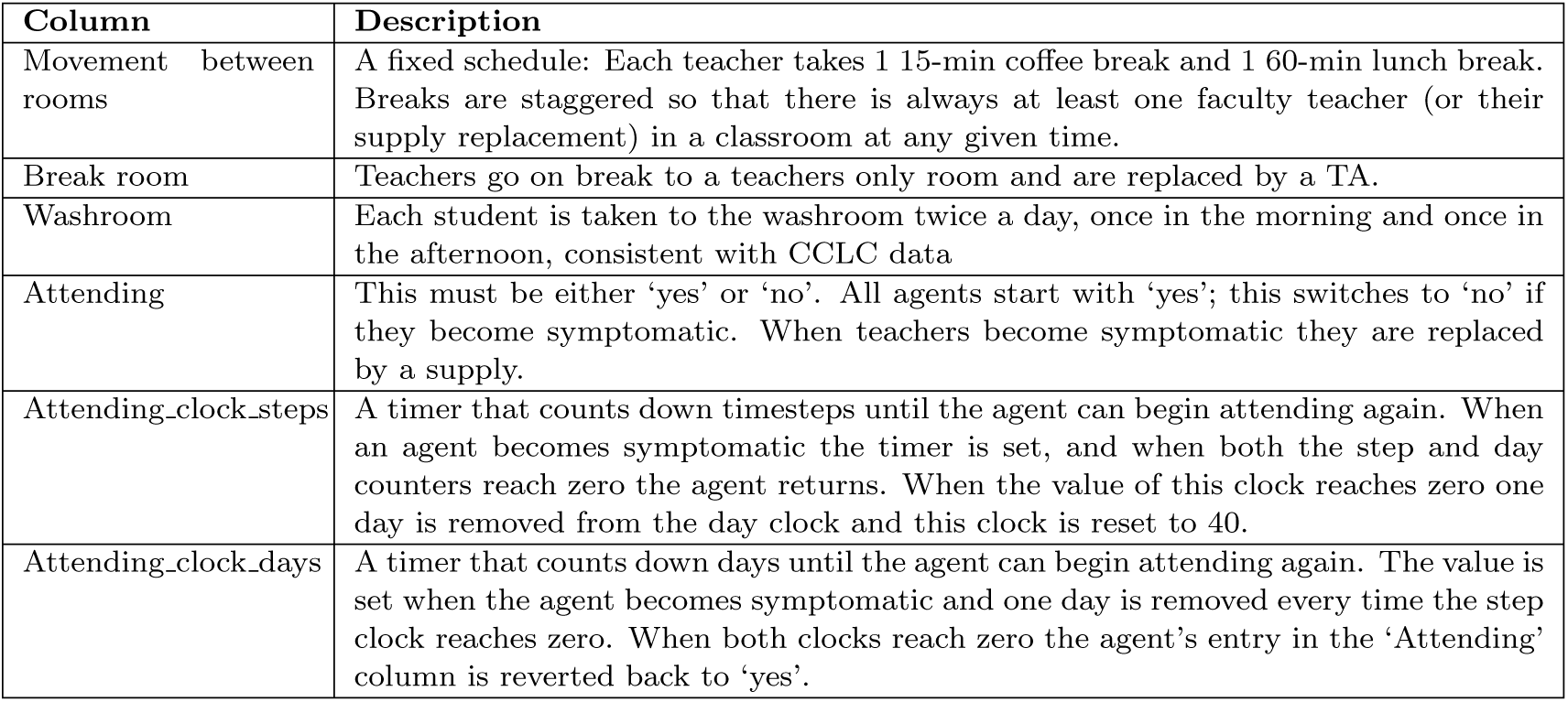
Default Columns.

| Column | Description |
| --- | --- |
| Type | Student or Teacher |
| Subtype | Student: toddler or preschooler. Teacher: regular, supply or teaching assistant |
| Arrival | Teachers always arrive at timestep 1, representing a 7:30 am start. Students arrive at a randomly determined timestep between steps 1 and 9 (7:30am to 9:45am). Note that student arrival times are determined once at the beginning of the simulation and remain the same throughout. |
| Departure | Teachers depart at timestep 40 (5:30 pm) unless they are sent home early due to being symptomatic. Students depart at a randomly selected time between time steps 31 and 40 (3:30pm to 5:30pm). Note that student departure times are determined once at the beginning of the simulation and remain the same throughout. |
| Agent's homeroom | This determines where agents go when they arrive at the facility. Although there is some movement between rooms each individual agent will spend the majority of their time in their home room. |
| Agent identifier | A numerical identifier unique to that agent *in their home room* (i.e. each name gets used once per homeroom).<sup>1</sup> |
| Room | Room the agent is currently in. If the agent is not at the facility this entry will be 'abs' for absent. |
| Movement between rooms | A fixed schedule: Each teacher takes 1 15-min coffee break and 1 60-min lunch break. Breaks are staggered so that there is always at least one faculty teacher (or their supply replacement) in a classroom at any given time. |
| Break room | Teachers go on break to a teachers only room and are replaced by a TA. |
| Washroom | Each student is taken to the washroom twice a day, once in the morning and once in the afternoon, consistent with CCLC data |
| Attending | This must be either 'yes' or 'no'. All agents start with 'yes'; this switches to 'no' if they become symptomatic. When teachers become symptomatic they are replaced by a supply. |
| Attending_clock_steps | A timer that counts down timesteps until the agent can begin attending again. When an agent becomes symptomatic the timer is set, and when both the step and day counters reach zero the agent returns. When the value of this clock reaches zero one day is removed from the day clock and this clock is reset to 40. |
| Attending_clock_days | A timer that counts down days until the agent can begin attending again. The value is set when the agent becomes symptomatic and one day is removed every time the step clock reaches zero. When both clocks reach zero the agent's entry in the 'Attending' column is reverted back to 'yes'. |

In general we number pathogens from {1, 2*, …, n*}, with *i* being the index of a pathogen. Six additional columns are added for each pathogen being simulated, and relate to an agent’s status with respect to that specific pathogen. These are described in Table 3

**Table 3.** Pathogen-Specific Columns.

|  |  |
| --- | --- |
| status_pathogen.i | The status of the agent with respect to pathogen i. Must be one of either 'susceptible', 'exposed', or 'infected'. |
| symptoms_pathogen.i | is the agent symptomatic with respect to pathogen i? |
| recovered_pathogen.i | Has the agent previously been infected by pathogen i? Must be either 'yes' or 'no'. |
| vaccinated_pathogen.i | Has the agent been vaccinated for pathogen i? Must be either 'yes' or 'no'. |
| pathogen_clock_steps.i | Used in conjunction with pathogen_clock_days.i to control the timing of transitions from 'exposed' status to 'infected' status and from 'infected' status to 'susceptible' with the recovered value set to 'yes'. |
| pathogen_clock_days.i | Used in conjunction with pathogen_clock_steps.i to control the timing of transitions from 'exposed' status to 'infected' status and from 'infected' status to 'susceptible' with the recovered value set to 'yes'. |

During each 15 minute timestep, a contact matrix is produced for every room in the simulation and these contact matrices are used to compute the probability that each susceptible agent in the room will gain the ‘exposed’ status based on their potential contact with an infected agent. To compute this probability we assume that, for any given contact between an infected agent *j* and a susceptible agent *k*, there is a probability, *P_jk_*, of the susceptible agent becoming exposed. The exact value of *P_jk_* will change depending on the type of agents in question, the pathogen in question, as well as the ‘recovered’ and ‘vaccinated’ status of the susceptible agent. The probability that susceptible agent *j* will *not* be exposed by infected agent *k* is (1 − *P_jk_*)*^n^jk* where *n_jk_* is the number of contacts between *j* and *k* in the given timestep. If we let *K* denote the set of all infected agents in the room then the totalITprobability that agent *j* will be exposed by interacting with all of them is 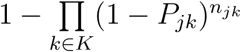.

When an agent is first exposed to a pathogen they are assigned a duration, specified in days and steps, until their status changes to ‘infected’. Once their status changes to ‘infected’ they are assigned a second duration, also in days and steps, which dictates how long they must wait until their status reverts to ‘susceptible’ and their recovered status changes to ‘yes’. Both the timing of the exposed period and the infected period are handled by *pathogen clock steps t* and *pathogen clock days t*, where *t* is the time index. At the same time that an agent’s status is changed from exposed to infected, a random number generator is used to determine if they are symptomatic or not. The probability of being symptomatic varies with respect to the agent type and the specific pathogen in question. If the newly infected agent is symptomatic, they are sent home and the attending clock is used to determine when they return. If the agent happens to be a teacher, they are replaced by a supply at this time as well.

Any time a clock is set with a specific duration, the duration is drawn from a range of possible values. For the duration of the exposed and infected periods the allowed range depends on the specific pathogen in question as well as the agent type. In the case of agents being sent home when symptomatic, the allowed range depends only on the agent type. Table 4 shows the likelihood of an agent (student or teacher) being absent from the CCLC for a number of days between 1 and 5.

**Table 4.** Sick-leave information upon infection with pathogens[10], [11].

| Duration of sick leave range, $t \in (1, \dots, 5)$ | 1 day | 2 days | 3 days | 4 days | 5 days |
| --- | --- | --- | --- | --- | --- |
| Relative likelihood of a symptomatic student to be absent on $t^{th}$ day | 25% | 30% | 20% | 15% | 10% |
| Relative likelihood of a symptomatic teacher to be absent on $t^{th}$ day | 10% | 15% | 20% | 30% | 25% |

To complete a simulation, it is necessary to provide parameters to specify each unique pathogen. Table 5 gives the names and descriptions of these parameters and their values for influenza and RSV. Besides student and teacher vaccination probabilities which can be omitted when there is no vaccination for that population for that month, the remaining parameters are essential to successfully completing a simulation. Two of the critical parameters, the probability of disease transmission per effective contact within CCLC on day *t* (pt sus *t*), and the probability that an agent is infected outside of CCLC on day *t* (offsite infection *t*), are unknown. In the next section, we estimate these values by numerical calibration, using the past known burden of disease and vaccine uptake in the population of the Wellington-Dufferin-Guelph public health region. *V_eff_* in the table represents the efficacy of an available vaccine.

**Table 5.**
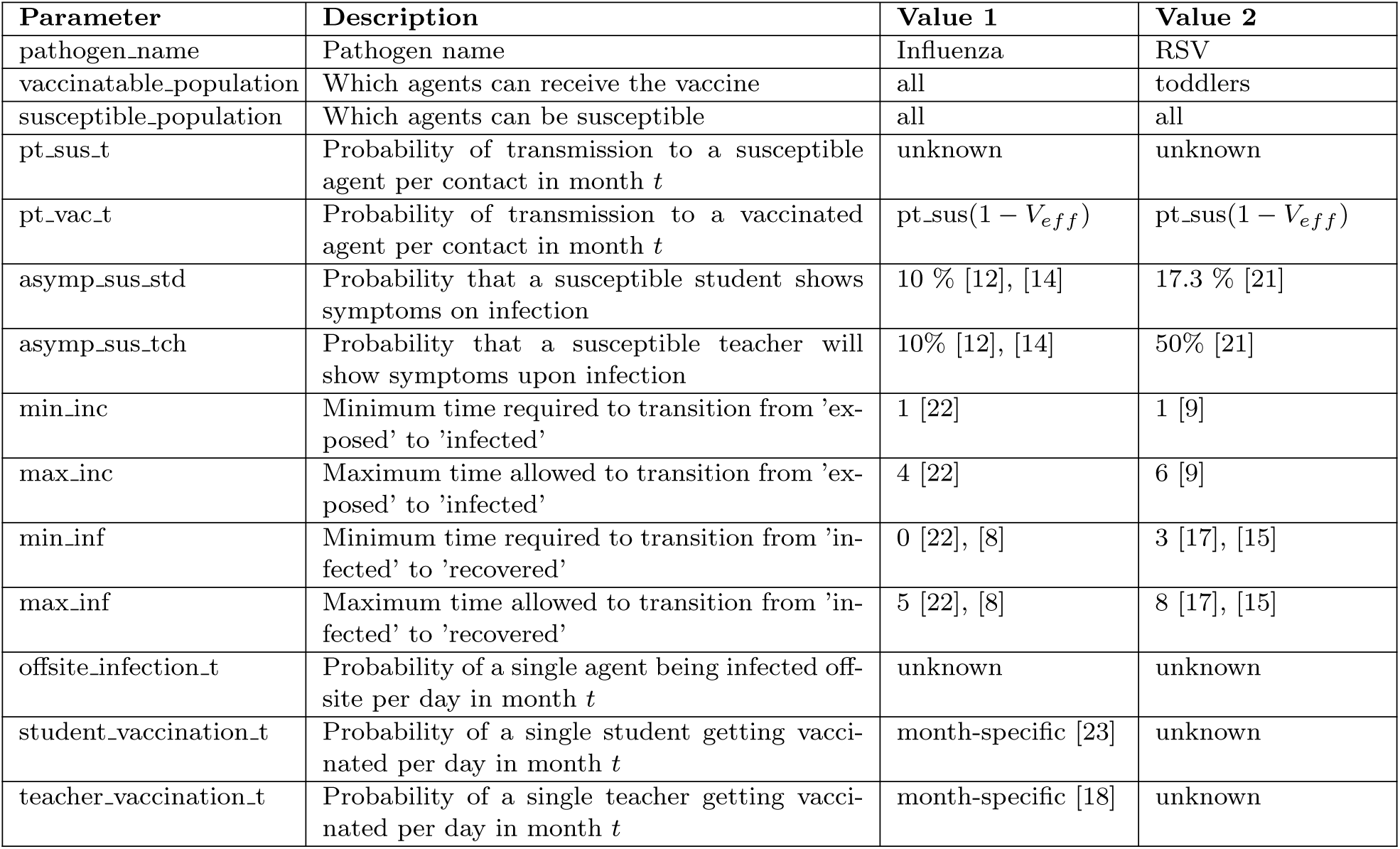
Transmission, vaccine and symptoms parameters per agent types.

| Parameter | Description | Value 1 | Value 2 |
| --- | --- | --- | --- |
| pathogen_name | Pathogen name | Influenza | RSV |
| vaccinatable_population | Which agents can receive the vaccine | all | toddlers |
| susceptible_population | Which agents can be susceptible | all | all |
| pt_sus_t | Probability of transmission to a susceptible agent per contact in month $t$ | unknown | unknown |
| pt_vac_t | Probability of transmission to a vaccinated agent per contact in month $t$ | $pt\_sus(1 - V_{eff})$ | $pt\_sus(1 - V_{eff})$ |
| asympt_sus_std | Probability that a susceptible student shows symptoms on infection | 10 % [12], [14] | 17.3 % [21] |
| asympt_sus_tch | Probability that a susceptible teacher will show symptoms upon infection | 10% [12], [14] | 50% [21] |
| min_inc | Minimum time required to transition from 'exposed' to 'infected' | 1 [22] | 1 [9] |
| max_inc | Maximum time allowed to transition from 'exposed' to 'infected' | 4 [22] | 6 [9] |
| min_inf | Minimum time required to transition from 'infected' to 'recovered' | 0 [22], [8] | 3 [17], [15] |
| max_inf | Maximum time allowed to transition from 'infected' to 'recovered' | 5 [22], [8] | 8 [17], [15] |
| offsite_infection_t | Probability of a single agent being infected off-site per day in month $t$ | unknown | unknown |
| student_vaccination_t | Probability of a single student getting vaccinated per day in month $t$ | month-specific [23] | unknown |
| teacher_vaccination_t | Probability of a single teacher getting vaccinated per day in month $t$ | month-specific [18] | unknown |

## 3. Estimating probability of infection per contact in the day care

The number of symptomatic influenza cases in Canada was estimated to be higher than laboratory confirmed cases by an expansion factor of 195.1 and 252.9 in the 2011-12 and 2012-13 influenza seasons respectively [24]. We apply an expansion factor of 200 to laboratory confirmed influenza cases per month in the Wellington-Dufferin-Guelph (WDG) public health unit in the 2023-24 season [25] and we numerically estimate the number of infections arising in the child care facility each month (October to March) by using estimated monthly attack rates. We assume that each person is infected once over the season [26]. The results are displayed in Figure 1.

**Figure 1.**
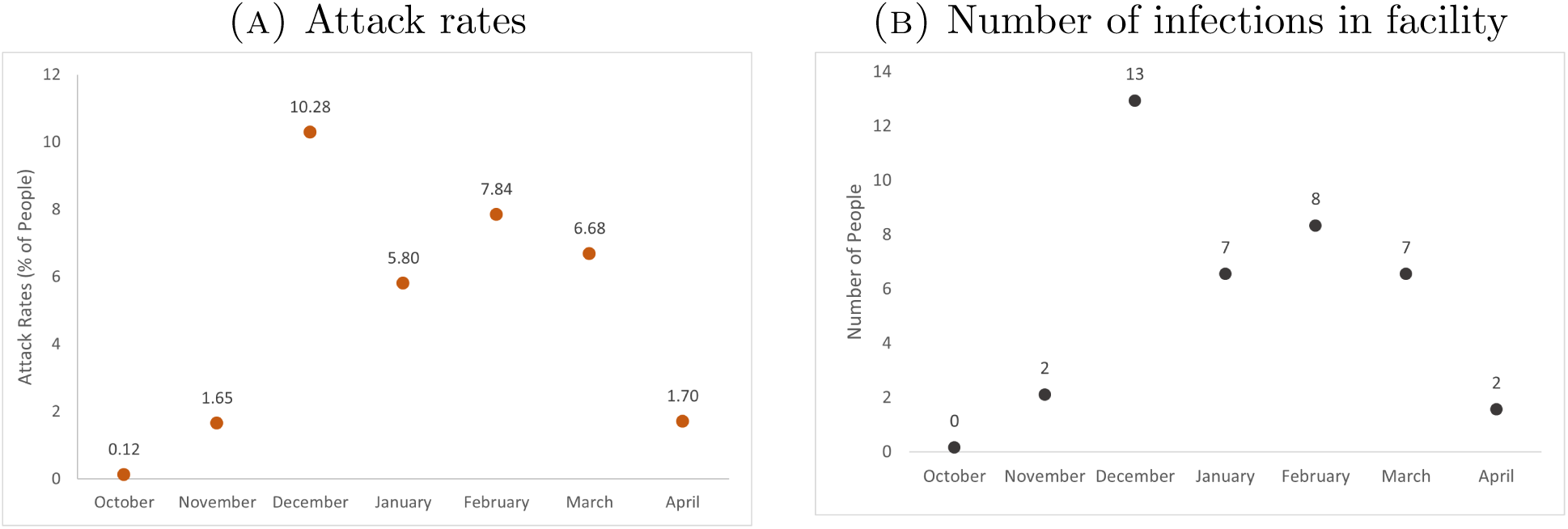
Seasonal pattern of influenza cases.

Given the number of expected infections at the facility, we want to find the probability of transmission pt sus *t* within the facility and the probability of a single agent being infected offsite in each month (offsite infection *t*). To do this, from the Latin Hypercube Sample package in *R*, we generate hundred random pairs of the two parameters in the interval (0, 1) for each month, ten times. Simulations are completed with each sample pair. Results with the number of infections approximately equal to the estimated number of infected from past data are recorded as fit input values.

We obtained the average values for the parameters each month by weighting each interval (0-0.1, 0.1-0.2, …) with the formula, 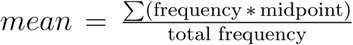. This yields a single representative value that reflects the combined contribution of all intervals and their respective frequencies. The process of: i) generating random pairs of parameter samples; ii) identifying desired parameter values corresponding to the infected numbers monthly data after a thousand simulations; iii) and finding the mean is repeated four, seven and ten times. In Figure 2, the range of values for each month is shown to lie within a small interval irrespective of the increasing number of iterations. The median values from 10 iterations will be used in generating results for the remainder of the paper. The vaccine efficacy of an influenza vaccine is taken to be *V_eff_* = 49% (see [27]).

**Figure 2.**
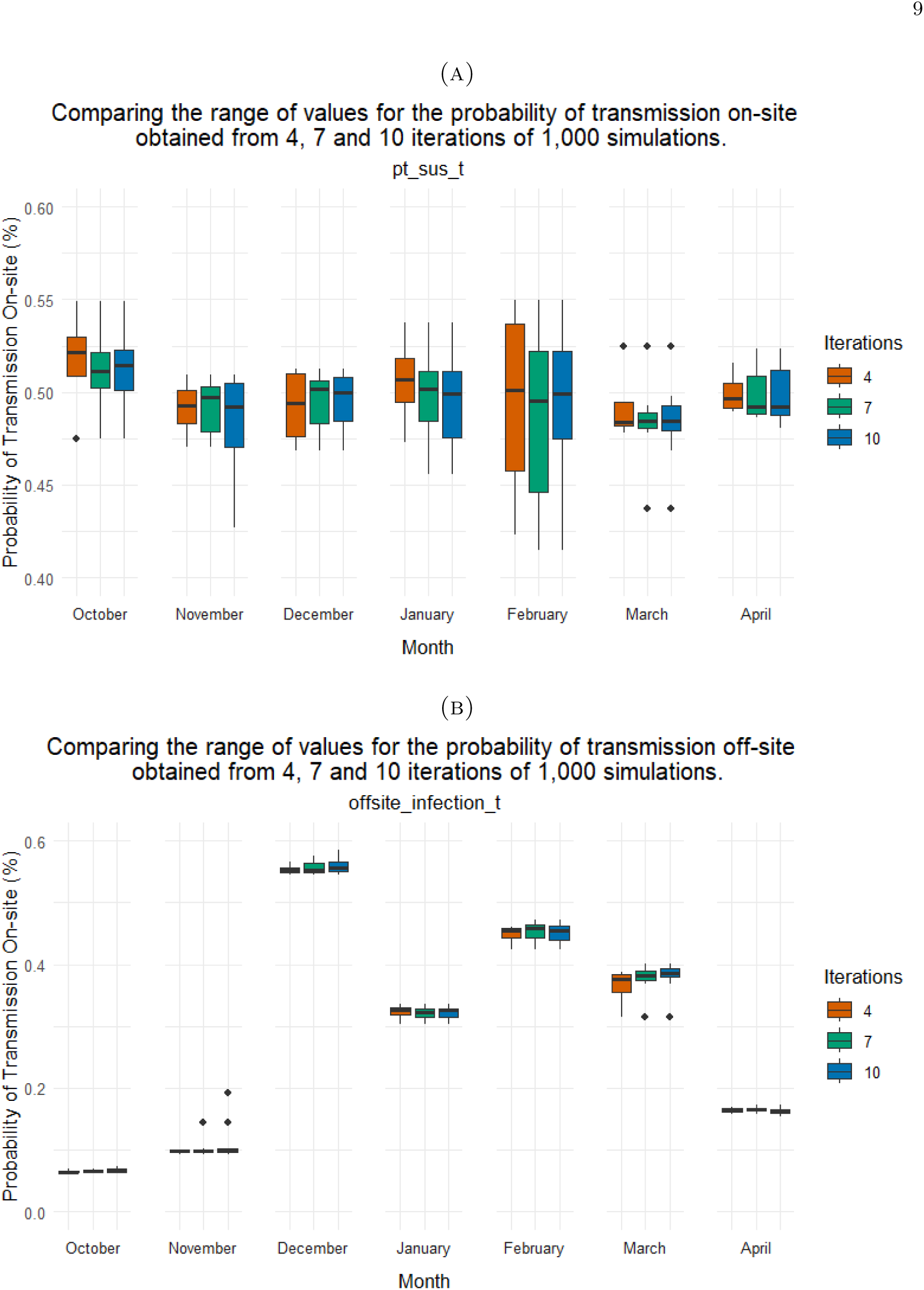
Boxplots showing estimates of the probability of disease transmission and the probability that an agent is infected outside the facility in a single day.

The median values are plotted in Figure 3. We observe that pt sus remains fairly stable between 0.48% and 0.52%. In the beginning of the season (October), the probability that an agent is infected outside the facility is very low. This increases in November and peaks in December, consistent with the general behaviour of the influenza season. In January, the decreasing number of infections in the data is reflected by a decrease in offsite infection probability. Towards the end of the season, we observe a second, smaller peak in offsite infection probability. Whilst the probability decreases in April, it remains higher than in the beginning of the season. This is because the majority of agents are vaccinated/recovered, and so are less/no longer susceptible so there is a smaller susceptible population of agents.

**Figure 3.**
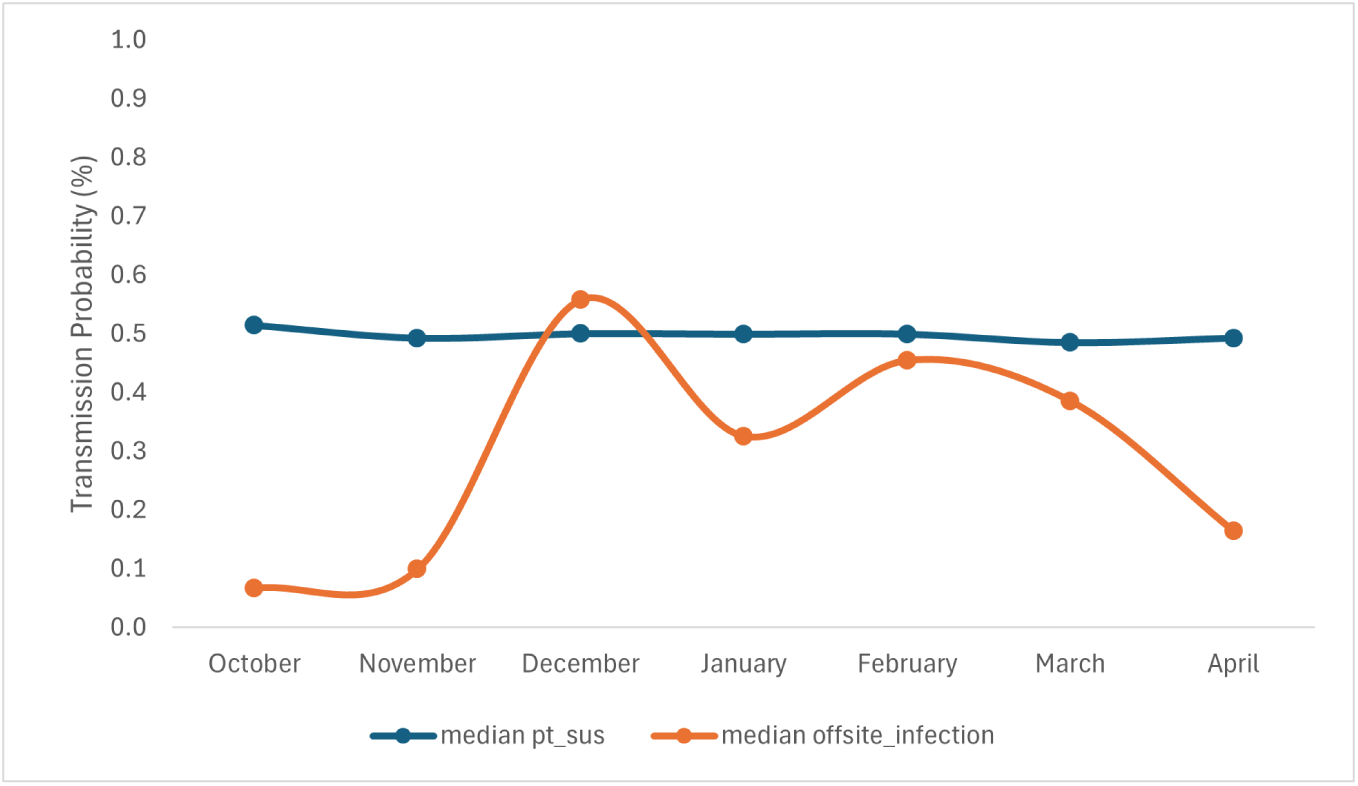
Median probabilities of influenza transmission within and outside the facility.

In 2023 approximately 44.5% of kids under 4 were vaccinated against flu in the Ontario region [23]. Given that we have 104 students in the learning center, we estimate that 45 - 50 students of the CCLC would be vaccinated against influenza. For adults, we estimate that 9-10 of the 24 adults in the center, i.e 38% [28] are vaccinated. An estimate of the percentage of students and teachers vaccinated at the facility each month is shown in Figure 4. We also assume that each person is vaccinated only once over a season [18]. Assuming that each month has 30 days and vaccinations are uniformly distributed throughout the month, we obtain the approximate daily vaccination probability for an individual.

**Figure 4.**
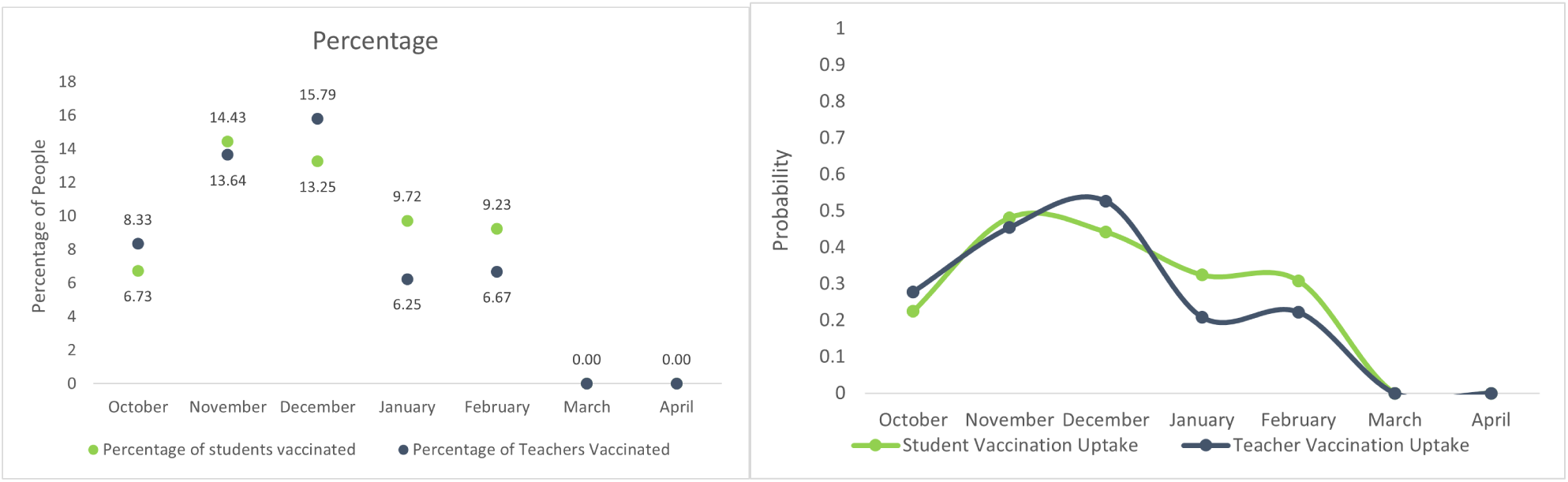
Percentage of people (students and teachers (left panel)). Estimated influenza vaccination probability per day (right panel).

Using the values of pt sus, offsite infection and vaccination probabilities that we have derived from our analyses, the median burden of disease among students for ten, twenty and thirty simulations are found to identify the number of simulations required for stable results. The results (Figure 5) display the median line and 7-day increments (points) across the season. We observe little variance in the results for different number of simulations. Subsequent results will be generated from twenty simulations.

**Figure 5.**
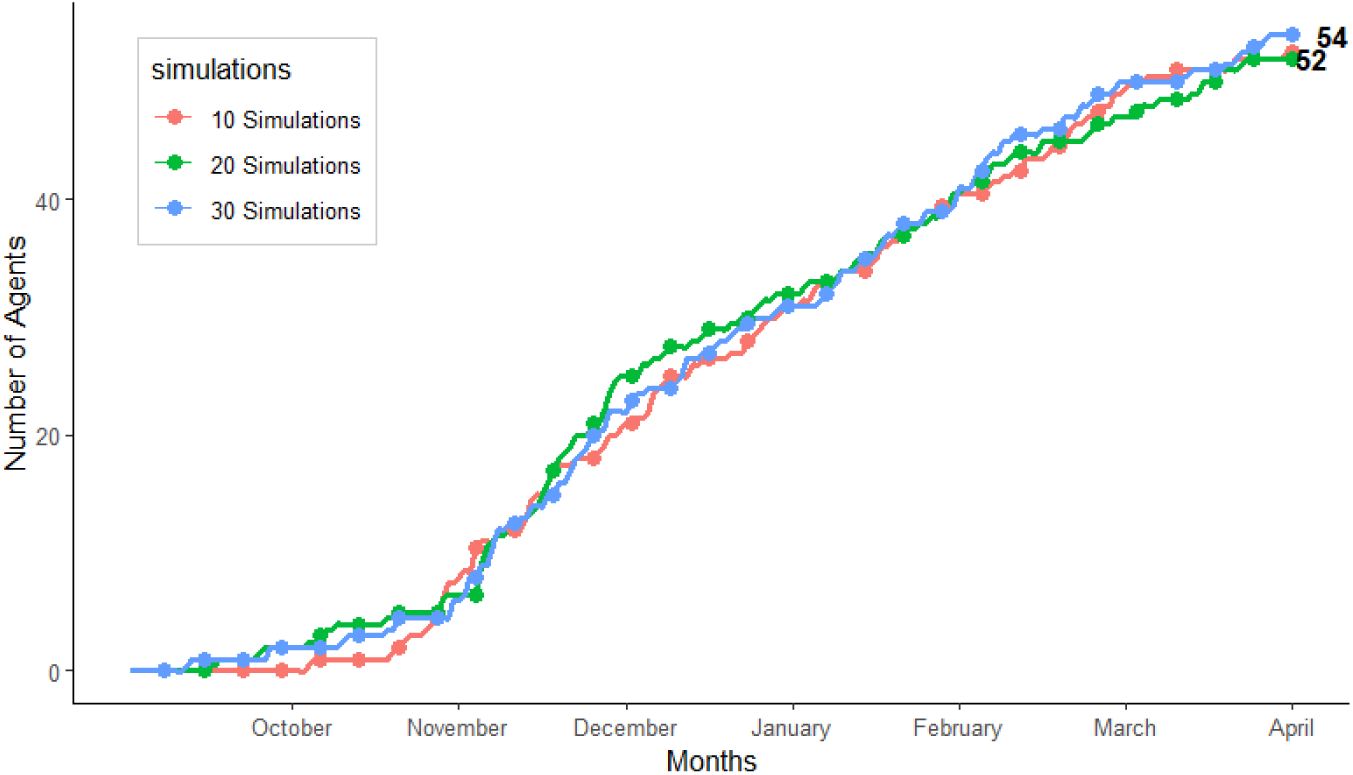
Burden of influenza among students for different number of simulations.

Figure 6 below displays the weekly trend of infections among students and teachers. Each image shows the median line and the 25th and 75th percentile ribbon. The visible points in the two lower panels are the 7-day cumulative values.

**Figure 6.**
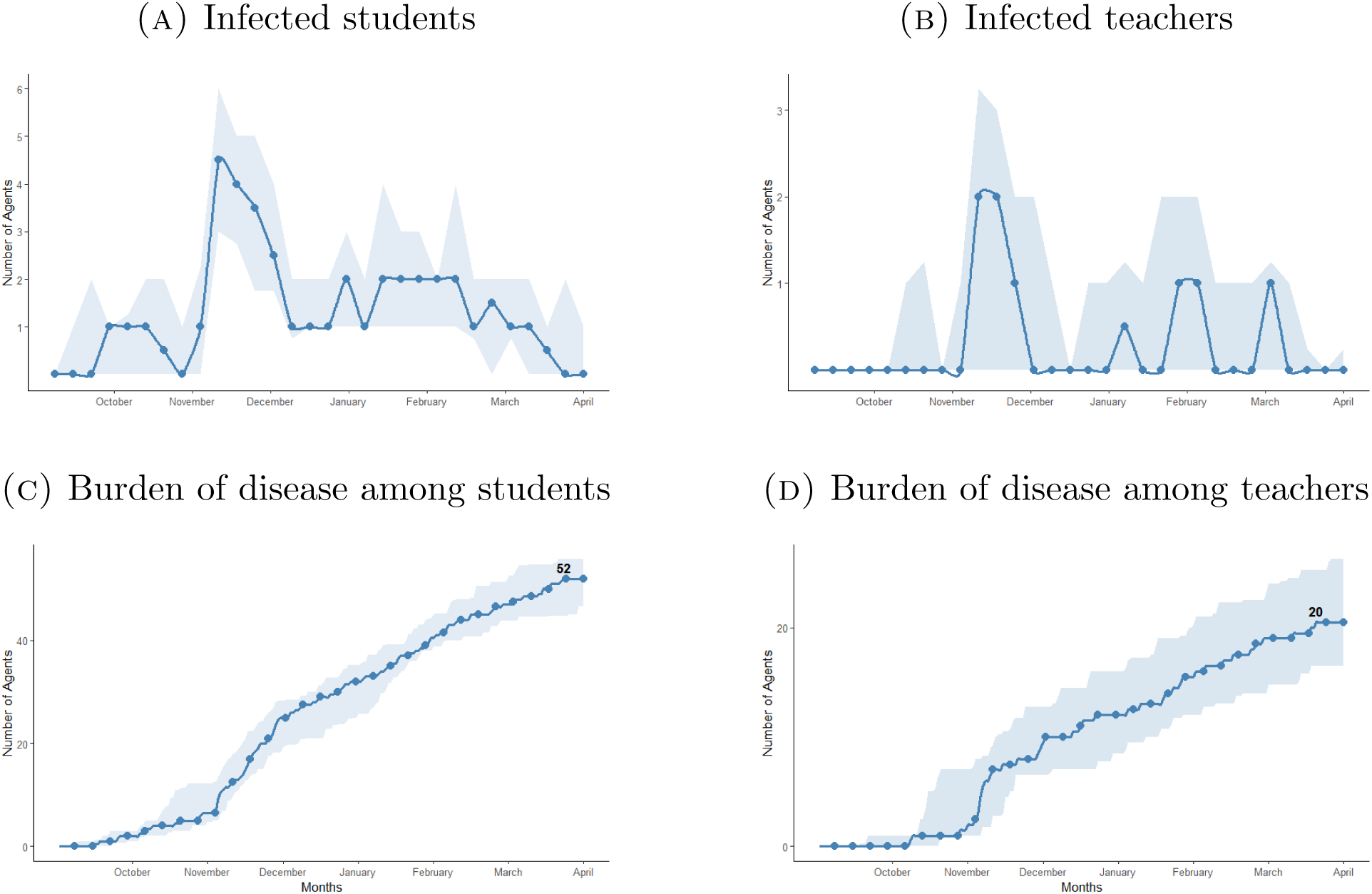
Results of simulation with influenza pathogen.

Consequently, we are able to estimate the likely number of absent students and teachers (Figure 7), and we can trace the vaccine uptake in the facility (Figure 8) within the two populations.

**Figure 7.**
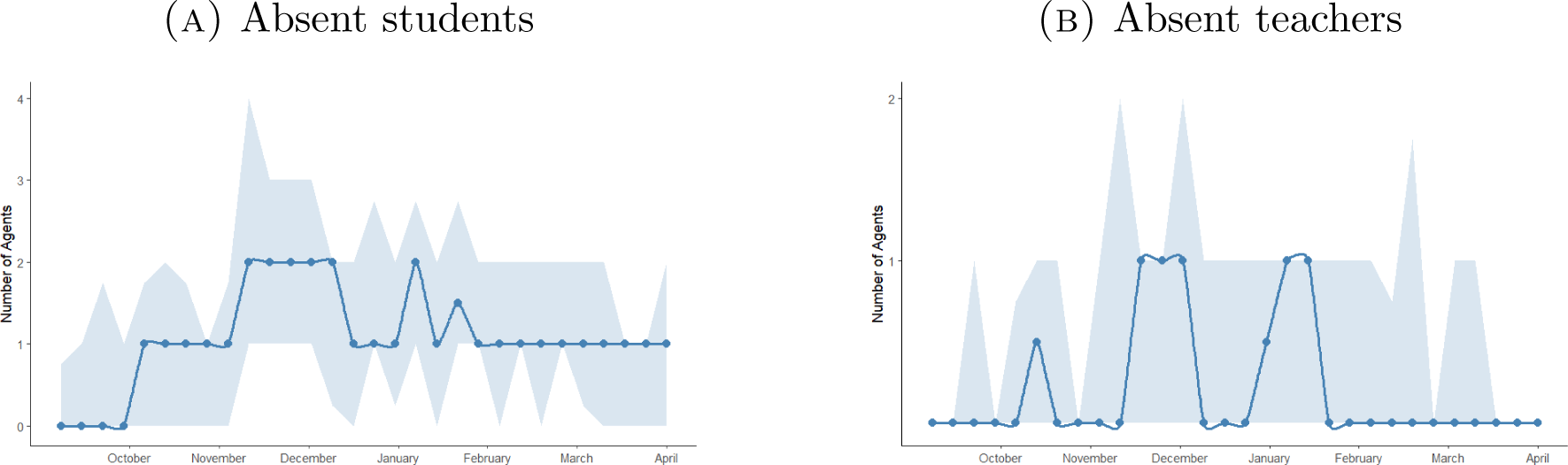
Absenteeism due to Vaccine uptake for the influenza pathogen.

**Figure 8.**
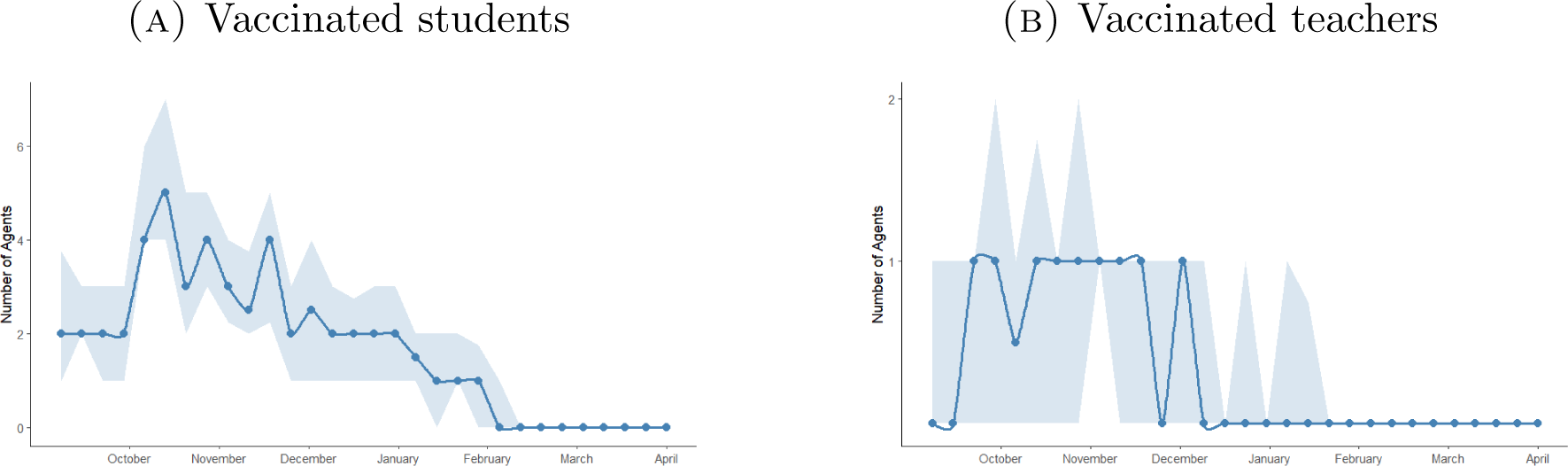
Vaccine uptake for the influenza pathogen in the facility.

## 4. Concurrent pathogens: influenza and rsv

In this section we run scenarios where two pathogens are spread concurrently in the facility, to draw us closer to real-life seasonal scenarios. Before we present our results, we discuss the relevant parameters for RSV, giving literature sources or mathematical approximations, as appropriate.

### 4.1. Deriving Unknown Parameters for RSV

We extend our results by deriving the probabilities of RSV infection in the facility and from offsite (Table 5) given the results we have obtained for flu. Let *R*_0_, *β*, *τ* and *γ* represent the basic reproduction number, the rate of disease transmission per effective contact, the rate of transition from exposed to infected and the recovery rate, respectively, as commonly denoted in an SEIRV compartmental model [29]. As it is widely recognized in the literature, [30]

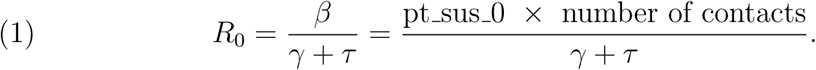

Assuming *R*_0_ ≈ 1.5 [31] for influenza, considering our parameters for influenza in Table 5 and pt sus t=0 = 0.5143 (for October, Figure 3), we have that 0.5143 × average no. of contacts

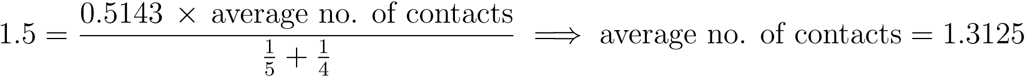

Applying now (1) for RSV, assuming a value of *R*_0_ ≈ 3 ([32]) and the same number of average contact numbers as above, we obtain: the probability of RSV infection per contact in the daycare at the beginning of the season (pt sus rsv0) from the equation

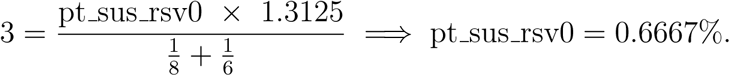

The results from simulations that apply our estimated pt sus rsv and offsite infection rsv values for RSV are shown in 9 (both RSV and influenza). As expected, we observe (Figure 9) a higher burden of disease for both students and teachers for the RSV pathogen.

**Figure 9.**
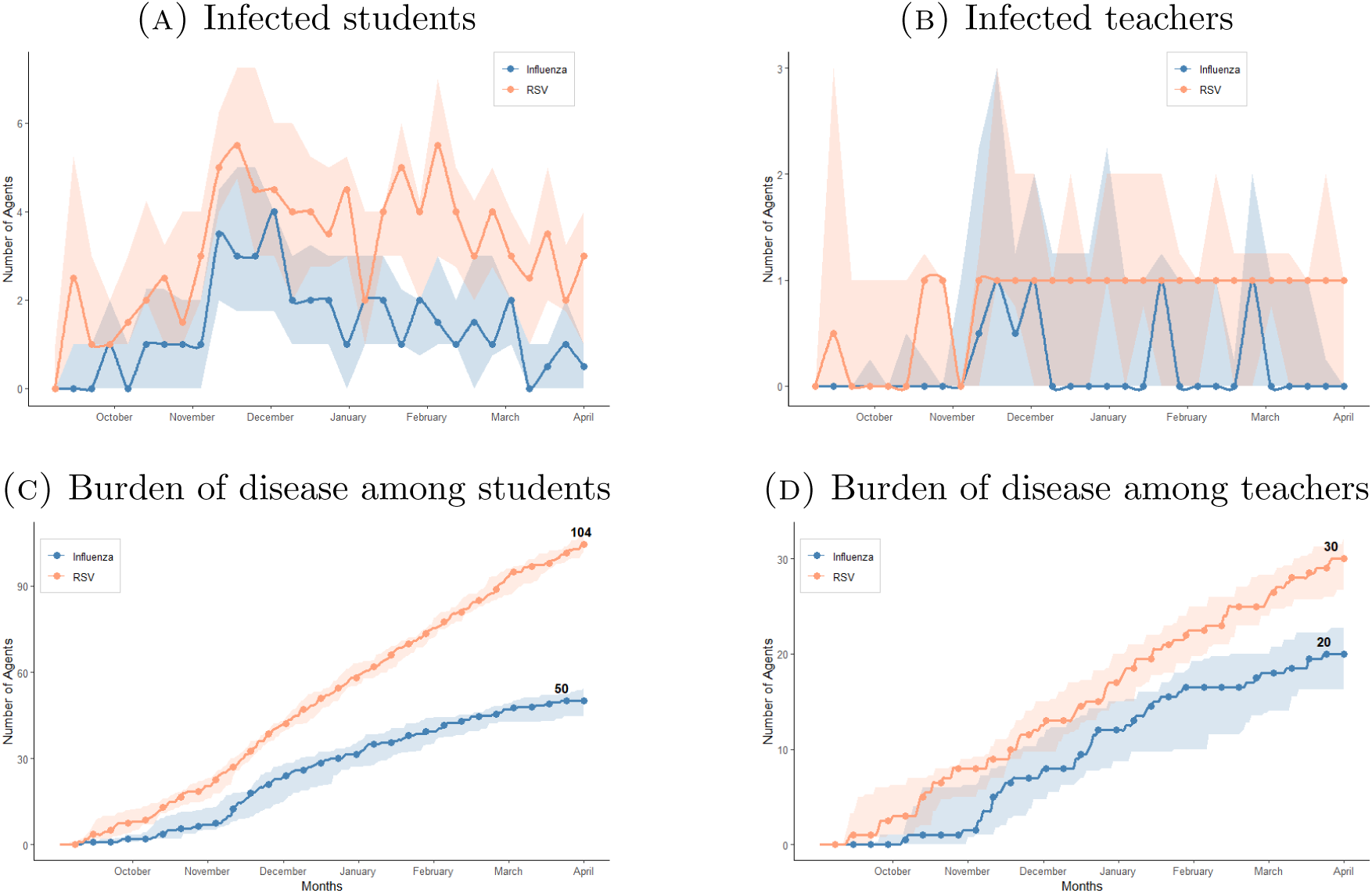
Results of simulation with both influenza and RSV pathogens. No vaccination for RSV.

## 5. Assessing RSV vaccination strategies in a small world environment

In this section we investigate the effect of vaccine availability on the burden of disease for RSV, varying vaccine efficacy and vaccination rates. We consider the scenario where an RSV vaccine is available for students only being available in the Fall of each year with a waning period of approximately five months. As adults are usually much less affected by contracting RSV, we will limit to presenting our results in the student population of the facility.

### 5.1. Assessing RSV vaccination impact across varying vaccination efficacy levels

We first assume that the student vaccination rates for RSV are the same as those used for influenza. We consider three scenarios, where the vaccine efficacy of the RSV vaccine is 50% (Figure 10), 75% (Figure 11) and 95% (Figure 12). For each scenario, both RSV and influenza pathogens are present.

**Figure 10.**
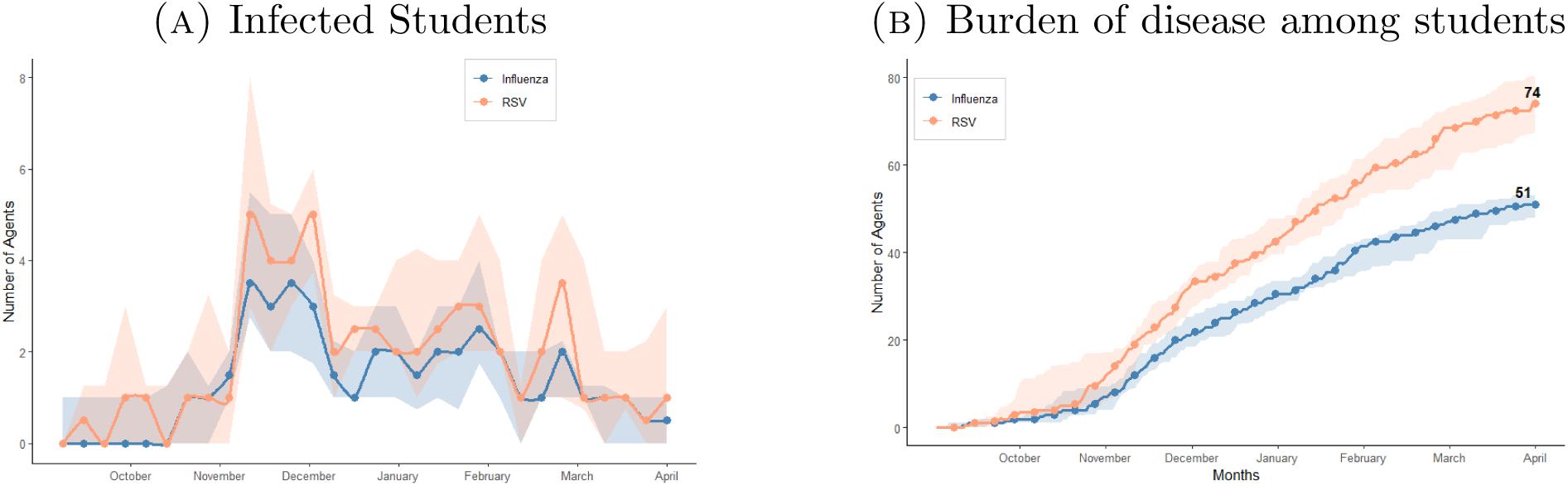
RSV vaccine with 50% efficacy.

**Figure 11.**
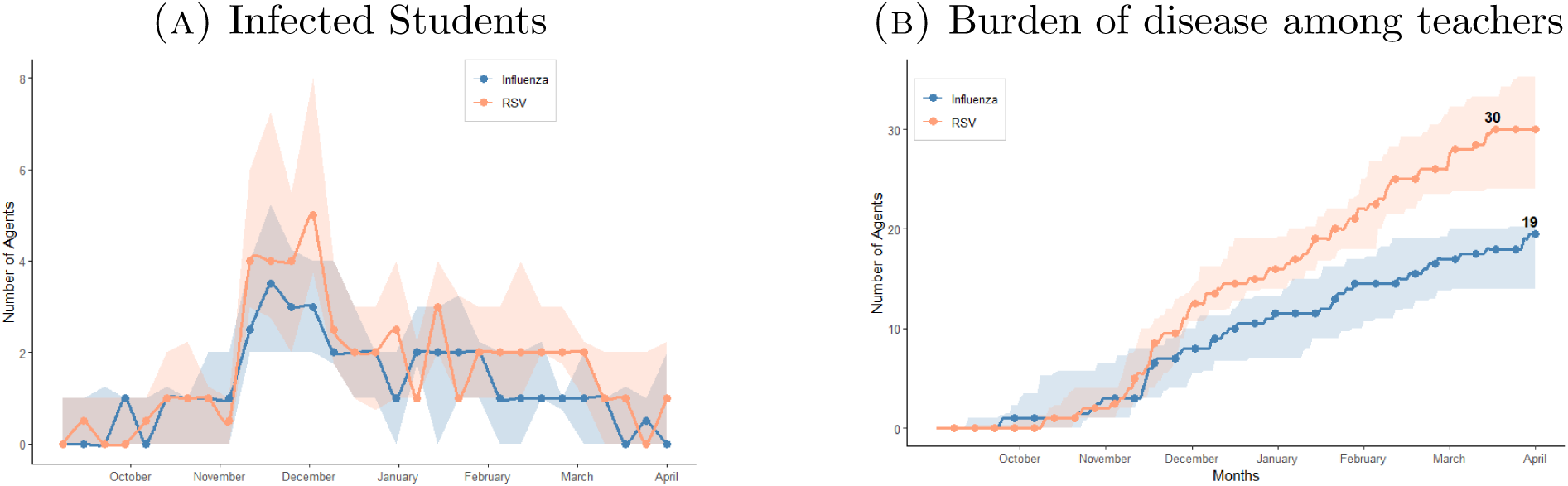
RSV vaccine with 75% efficacy.

**Figure 12.**
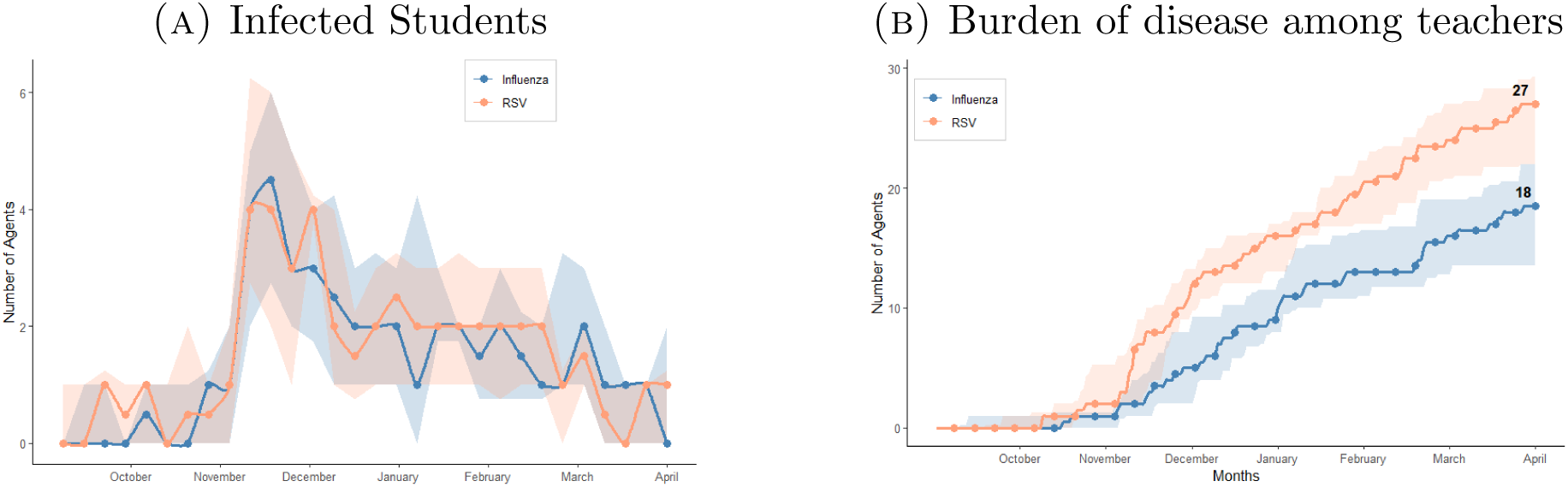
RSV vaccine with 95% efficacy.

In Figure 13, we isolate the changes in the burden of RSV disease among students for different vaccine efficacy. We observe that the initial introduction of a vaccine with an efficacy of 50% leads to a 30% decrease in disease burden. Subsequent increases by 50% (to 75%) and 27% (from 75% to 95%) in vaccine efficacy lead to an 16% and 15% decrease respectively in the burden of disease. This depicts a non-linear relationship between vaccine efficacy and the burden of disease. As efficacy increases, the burden of disease decreases but at a slower rate.

**Figure 13.**
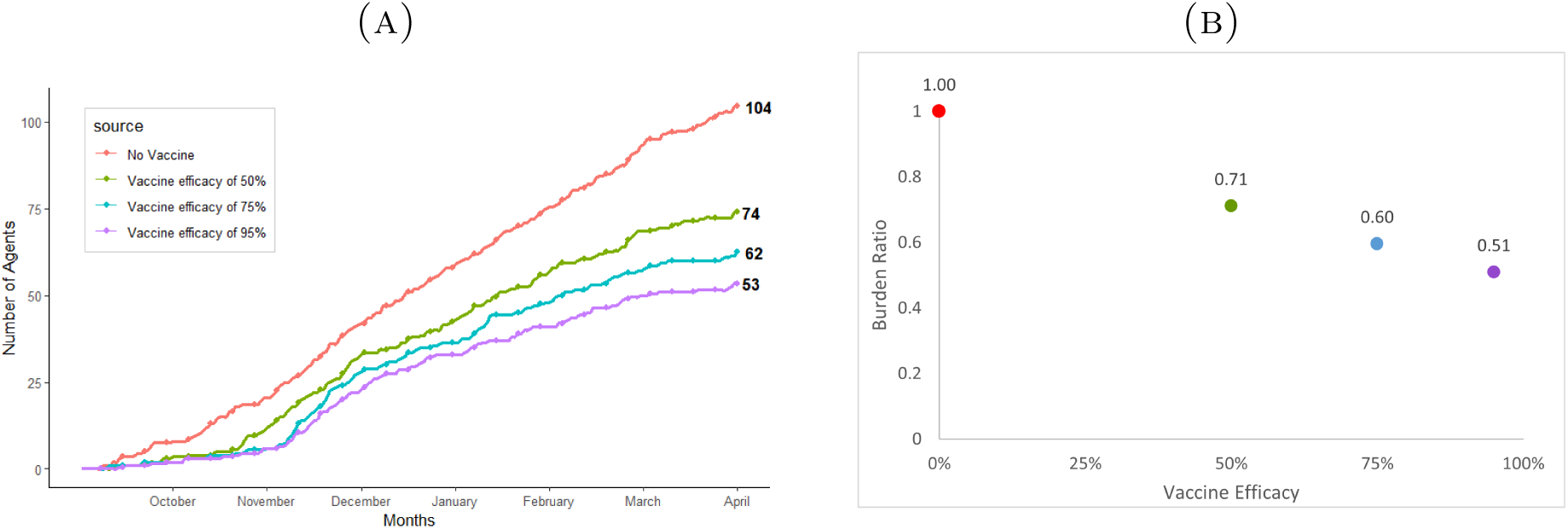
RSV Burden of disease on students for different RSV vaccine efficacy.

### 5.2. Assessing RSV vaccination impact across varying vaccination uptake levels

In this section, we compare the effect of changes in the daily vaccination probabilities for each month to the burden of disease among students. We increase the base vaccination probabilities (from influenza) Figure 4 by 0.50 (Figure 14), 0.75 (Figure 15 and 1 (double the probability, Figure 16). Vaccine efficacy is maintained at a constant 50%.

**Figure 14.**
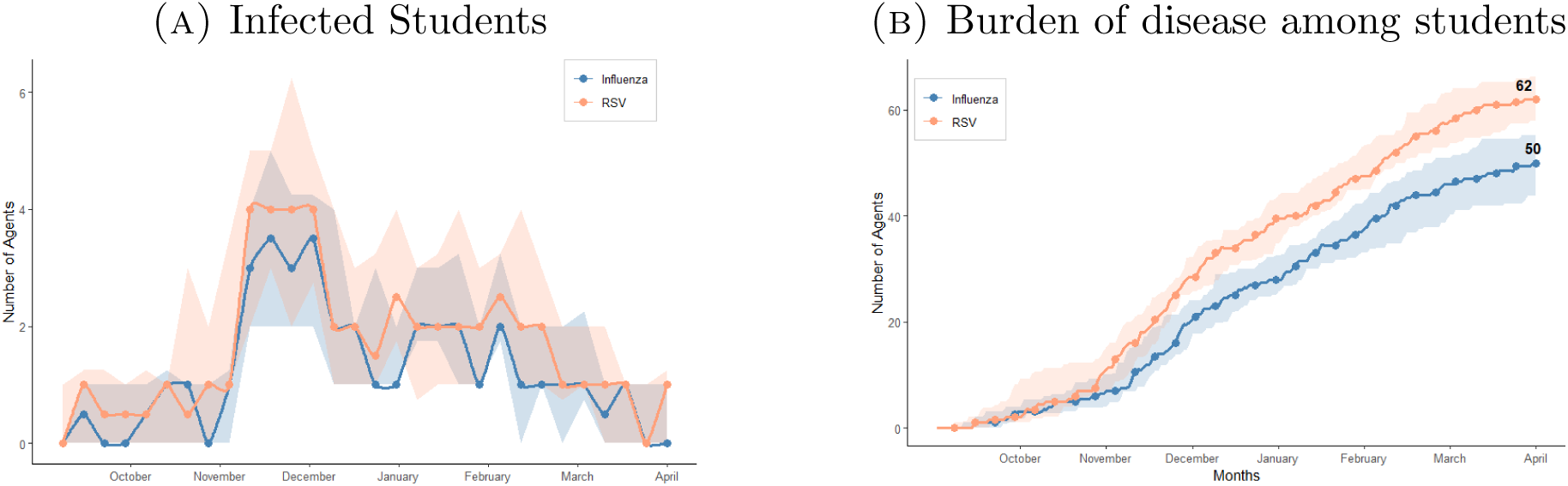
Considering RSV vaccine with 50% increase in vaccination probabilities.

**Figure 15.**
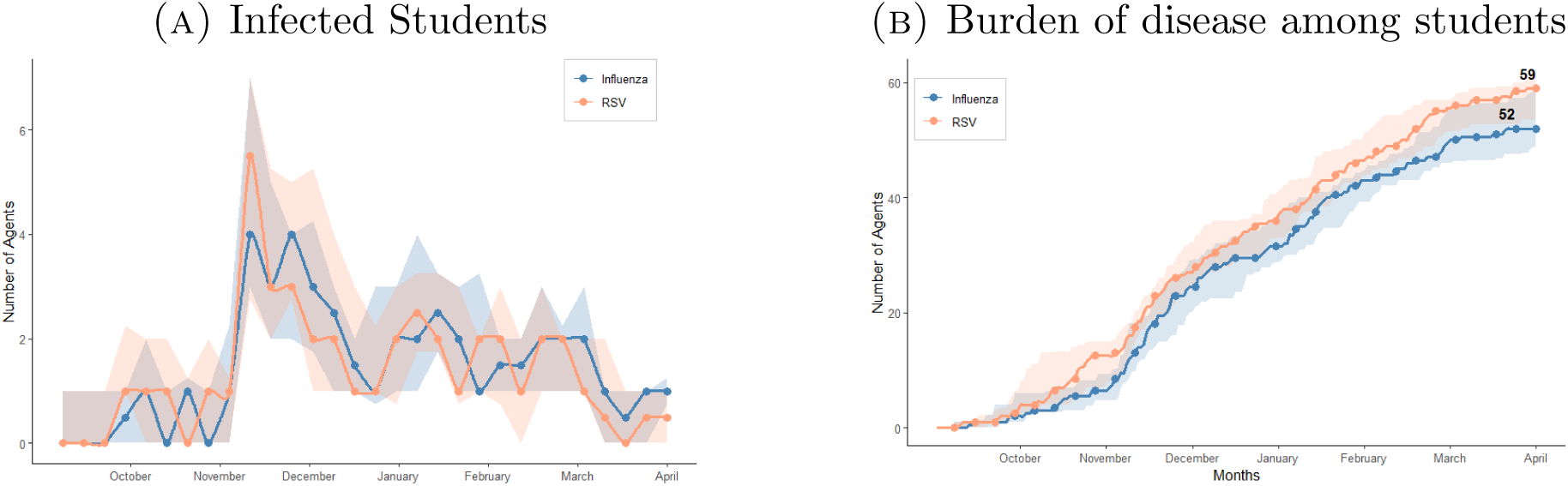
Considering RSV vaccine with 75% increase in vaccination probabilities.

**Figure 16.**
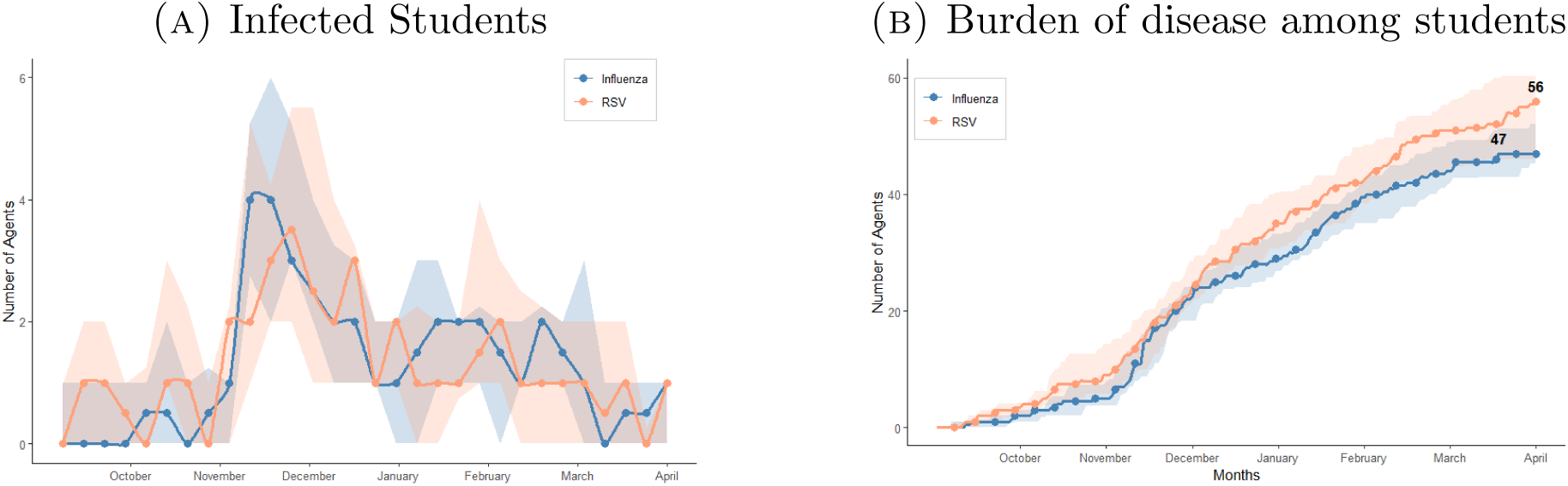
Considering RSV vaccine with double the initial vaccination probabilities.

For a vaccine with 50% efficacy, increasing the vaccination probabilities for each month has a more notable impact on the burden of disease of RSV compared to further increases in efficacy (Figure 17). Whilst the rate of decrease is non-linear as in the case of changes in efficacy, the decrease is faster.

**Figure 17.**
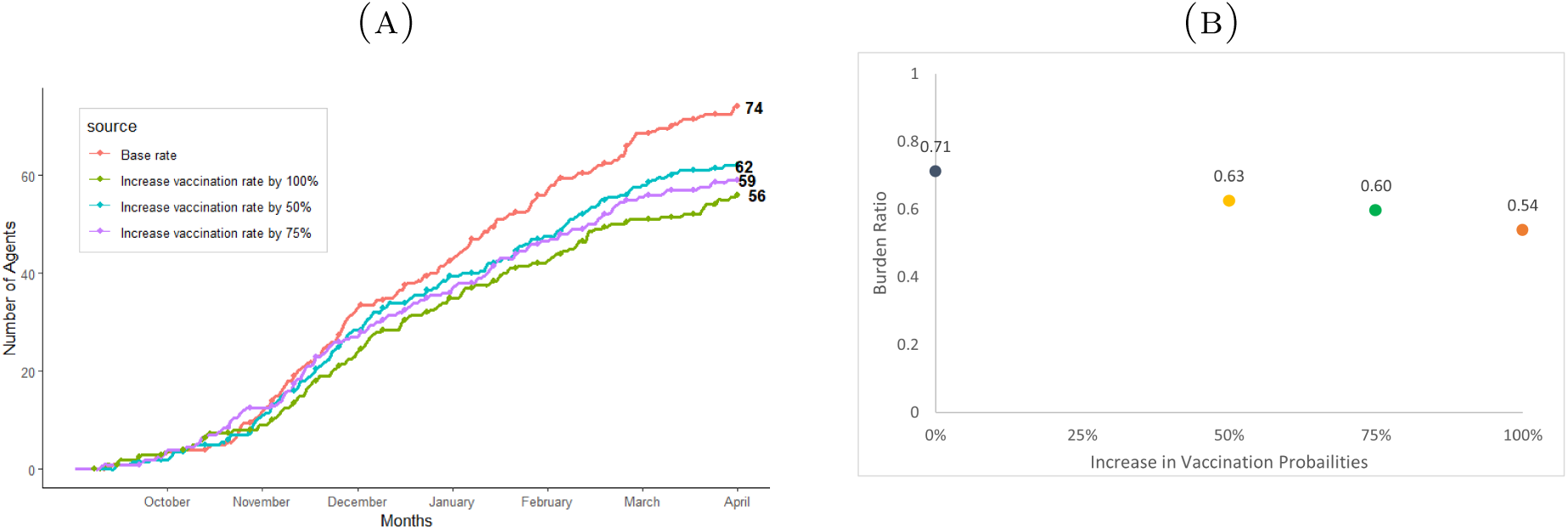
Comparing the burden of disease for RSV among students for varying vaccine probabilities and a constant efficacy of 50%.

## 6. Conclusion

Our simulations first established how influenza moves through a childcare facility, providing measured transmission rates that anchor the wider analysis. Building on that baseline, we turned to RSV and explored different vaccination strategies. Once vaccine efficacy reaches a moderate threshold, additional benefit comes from boosting uptake rather than focusing on marginal improvements in the vaccine itself. This insight shifts attention from trying to engineer a perfect vaccine to designing campaigns that reach more families. School leaders can use the projected infection curves to gauge the need for supply staff, anticipate student absences, and test different policies such as increased vaccination drives or stricter stay-home-when-ill rules before committing resources.

Linking each child to household contacts would capture how in-centre outbreaks spill over to older relatives (such as the aged, in the case of RSV). Scaling the contact network to several Ontario childcare centres would reveal how local epidemics merge or remain isolated, and the interplay with disease spread in the environment. Running the model across consecutive seasons could be used to investigate how partial immunity or waning protection could alter the seasonal behavior in the next season, or affect the effectiveness of control measures. Packaging these capabilities in a user-friendly dashboard would give school administrators and public-health officers a scenario lab for policy evaluation.

## Data Availability

All data produced in the present work are contained in the manuscript

